# Applying models of care for total hip and knee arthroplasty: External validation of predictive models to identify extended stay prior to lower-limb arthroplasty

**DOI:** 10.1101/2020.08.24.20180653

**Authors:** Meredith Harrison-Brown, Corey Scholes, Kam S. Sandhu, Milad Ebrahimi, Christopher Bell, Garry Kirwan

## Abstract

**Introduction/Aims:** Multiple screening tools exist for identifying patients at risk of extended stay following lower limb arthroplasty. Use of these models at other hospital sites requires verification of appropriate data coverage and evidence of validity in a new population. The aim of this study was to adapt and assess 1) data compatibility, 2) discrimination, and 3) calibration of three published models for identifying patients at risk of an extended (5+ day) stay, or those likely to stay for the target 3 or fewer days following lower limb arthroplasty.

**Methods:** Retrospective study, utilising a randomly selected (N=200 of a total 331 available in the electronic medical record) cohort of lower-limb Total Joint Arthroplasty (TJA) patients, to externally validate an adaptation of predictive tools and regression models published by three independent groups: Winemaker et al (2015)^1^, Oldmeadow et al (2003)^2^ and Gabriel et al (2018)^3^. Electronic medical records of a single, medium-sized public hospital were accessed to extract data required for the models and respective predictive tools, and model characteristics (included predictors, data coding, sample sizes) were modified according to the available data.

**Results:** The study cohort comprised 200 patients (60% female) at a median 70yrs of age (IQR 62-75). Approximately 58% received total knee arthroplasty (TKA) and 42% underwent total hip arthroplasty (THA). The two prediction tools and three regression models all required modifications due to data items being unavailable in the electronic records. A modification of the RAPT tool applied to 176 eligible patients resulted in sensitivity of 85.71% (95%CI 71.46-94.57) and poor specificity 32.09% (24.29-40.70), with 68% of short-stay patients classified in the high risk group. Adaptation of the second tool to 85 eligible patients resulted in unreliable estimates of sensitivity due to limited data. The three adapted regression models performed similarly well with regard to discrimination when used to predict patients staying for 5 days or longer (concordance index: Winemaker et al:, 0.79, n=198; Oldmeadow et al: 0.79, n=176), or those staying 3 days or less (Gabriel et al: 0.70, n=199). Estimates of calibration suggested the models were relatively well calibrated (spiegelhalter Z -0.01-0.29, p>0.05), although calibration plots indicated some variation remained unaccounted for, particularly with patients considered at ‘intermediate’ risk.

**Conclusion:** The three resulting regression models performed adequately in terms of discrimination and calibration for identification of patients at risk of an extended stay. However, comparison with published models was hampered by systemic issues with data compatibility. Further evaluation of such models in a specific hospital setting should incorporate improvements in data collection, and establish key thresholds for use in targeting resources to patients in need of greater support.

## Introduction

Although most patients undergoing lower limb arthroplasty can safely follow a short stay post-operative protocol, certain patients are at greater risk of complications or suboptimal clinical improvement necessitating an increased length of stay (LOS) in hospital ^4-9^. Identifying these patients prior to surgery would allow clinical teams to implement preventative steps and to allocate resources appropriately to each individual patient.

Over the last decade there has been a steady increase in total joint arthroplasty (TJA) putting an increased burden on healthcare costs. The Australian Orthopaedic Association National Joint Replacement Registry Annual Report (2019) ^10^ states there were 95 152 primary total hip and knee replacements performed in 2018, representing an increase of 3.4% from 2017 ^11^. Similar trends are noted across comparable health services, for example the number of joint replacements performed in Canada has increased over the past 5 years, with 130,000 surgeries now performed annually, with a resultant increase of 17.4% and 17%, respectively, over the last 5 years ^12^. Similarly, in the USA cumulative procedural volume has increased from 48,084 in 2012 to 1,525,435 in 2018 with primary knees (55.1%) and primary hips (33.1%) of the total during that time period ^13^. With current Australian population and TJA growth rates, the number of joint replacements is projected to undergo unsustainable 10-year growth of 276% and 208% for knee and hip replacements, respectively ^14^. Therefore the healthcare system is under significant pressure to reduce the costs associated with these procedures. Current evidence indicates a reduction in length of hospital stay is associated with a significant reduction in costs ^7,15,16^, therefore this metric has become a key target for improvement.

Available evidence has demonstrated an ability for some tools to assist in predicting length of stay and discharge location at certain sites ^1-3,5,9,17^. Within the clinical setting, such tools are being utilised, however, there is limited evidence to show that such tools can be applied indiscriminately across all clinical settings. Therefore, the aim of this study was to adapt and assess 1) data compatibility, 2) discrimination, and 3) calibration of three published models for identifying patients at risk of an extended (5+ day) stay, or those likely to stay for the target 3 or fewer days following lower limb arthroplasty.

## Methods

### Study design

This study is an adaptation and external validation of multiple predictive models with a randomly selected subset of medical records in a retrospective cohort design. The study was reported according to the RECORD guidelines for studies conducted using routinely collected health data ^18^ and the TRIPOD guidelines for reporting of prognostic models ^19^. In brief, the analysis flow commenced with a literature review to identify candidate tools or models already in existence in the literature, an assessment of predictive tools reported by two of the three included studies, followed by an assessment of the regression models from which the selected tools were derived, as well as the additional regression model reported by Winemaker et al ^1^. The present study is a Type 4 analysis as per published guidelines ^19^.

### Setting

The study is a single-centre analysis with 14 consultant surgeons operating within an orthopaedic department of a medium-size metropolitan public hospital located in South-East Queensland, Australia. Of the 200 randomly selected cases, a majority of the procedures were performed by 4 surgeons with 38, 34, 29 and 25 procedures respectively accounting for greater than 60% of surgeries. For the remaining TJA procedures, 4 surgeons accounted for 50 (25%) procedures with an additional 6 surgeons performing 10 or fewer procedures each. Data was collected as part of standard medical records compilation for lower limb total joint arthroplasty performed 2-Feb-16 to 4-Apr-19. Medical records were entered by hospital staff directly into the integrated electronic medical record (iEMR) (Cerner, USA) introduced to the public health system within the state as of Nov-2018 as part of a digital hospital rollout, with the specific hospital converted in Apr 2019. The iEMR records were accessed for data collection purposes between Sep-19 and Feb-20.

### Treatment details

All patients electing to undergo total knee or hip arthroplasty for end-stage arthritis underwent standard multidisciplinary preoperative screening incorporating anaesthetic, orthopaedic, nursing and allied health review to ensure fitness for surgery. All eligible patients were subject to a standardised ERAS protocol involving the use of spinal anaesthesia and tranexamic acid, chemical DVT prophylaxis (aspirin or low molecular weight heparin as appropriate) and attempted mobilisation on the day of surgery. Fitness for discharge was assessed on a daily basis, defined as functional safety (ability to mobilise safely) as determined by a qualified physiotherapist; and medically stable as determined by a medical practitioner. If the patient was identified by the physiotherapy team as requiring additional rehabilitation once declared medically stable, an orthogeriatric review was completed to enable referral to an external rehabilitation facility.

### Ethics and Governance

Ethical approval for the study was granted by the Metro South Human Research Ethics Committee (HREC/2019/QMS/57093); and access to the dataset was approved by the Metro South Health Research Governance Committee. Data was retrieved by hospital staff from the electronic medical record and de-identified data was then distributed to the research team for further research, analysis and modeling.

### Models/tools for validation

A literature search was conducted in Feb 2019 to identify a selection of potential models that may be appropriate for identifying patients at risk of extended stay following in-patient total knee arthroplasty.The three studies selected contained predictive models based on commonly collected preoperative factors including demographics, comorbidities, mobility, use of opioid painkillers, and select psychosocial factors such as availability of community support.

A brief summary of model characteristics is presented below, and a list of key outcomes and predictors is displayed in **Table 1**.

**Table 1:** Summary of models/tools assessed for validation

|  | **Winemaker et al 2015** | **Oldmeadow et al 2003** | **Gabriel et al 2018** | **Oldmeadow et al 2003 - RAPT** | **Gabriel et al Score** |
| --- | --- | --- | --- | --- | --- |
| **Final sample (N)** | 1459 | 520 (testing);<br>126 (validation) | 644 (testing);<br>316 (validation) |  |  |
| **LoS Outcome** | 3 or less; 4; 5 or greater | Discharge destination* | less than or equal to LOS of 3 days | Discharge home vs rehabilitation facility | less than or equal to LOS of 3 days |
| **Model/tool specifications** | generalized logistic regression model | Logistic regression model | multivariable logistic regression |  |  |
| **Final variables** | Gender; Joint; Age; ASA; No. cardiovascular comorbidities; No. of renal comorbidities; Current smoker; diabetes oral tx | Age; Gender; Mobility status; Gait aids; Community support; Caregiver | Age; Preoperative opioid use; Preoperative METs; Neuraxial anaesthesia; Gender; No preoperative anemia; No COPD; No hypertension; Non-obesity |  |  |

#### Prediction scoring tools

##### Tool 1

The first predictive tool ^2^ was the Risk Assessment and Prediction Tool (RAPT), a simple scoring tool used developed to predict the likelihood of a patient being transferred to an extended stay facility (rehabilitation). It is based on scores attributed to responses from six questions related to a patient’s demographics, walking capacity and living arrangements. The tool scoring system was developed from a logistic regression model with the odds ratios for significant predictors used to weight the scores given to each response. The RAPT uses thresholds applied to the total score to place a patient in a risk category associated with discharge to a facility; in the original cohort a score of 0-5 indicates high risk of transfer to inpatient rehabilitation, 6-9 indicates intermediate risk, and 10-12 indicates low risk.

##### Tool 2

The second predictive tool ^3^ was limited to total hip arthroplasty cases with a target category LoS of ≤3 days. The score was based on patient demographics, opioid usage and comorbidities. The tool scoring system was developed from a logistic regression model with the odds ratios for significant predictors used to weight the scores given to each response.

#### Regression models

##### Model 1

The first model ^1^ assessed in the present study used a generalized logistic regression model to fit to a three-level classification of length of stay, with ≤3 days used as the reference level, 4 days and ≥ 5 days being the remaining levels. A stepwise elimination approach was applied to a starting predictor list of 102 factors using an α of 0.2. The model was developed using SAS (v9.3, SAS Institute), R (v3.03, R Foundation, Austria) and SPSS (v20, IBM Corp, USA).

##### Model 2

In the event that the scoring tools did not perform well on the selected dataset the underlying model for Tool 1 ^2^ was also adapted to the selected dataset. The logistic regression model structure is not fully described in the original paper, so it was assumed that the model was based on a logit link function. The original model was developed in SPSS (v10, SPSS inc, USA).

##### Model 3

The model used to derive Tool 2 ^3^ was adapted to the selected dataset. Feature selection was performed through stepwise (forwards-backwards) elimination based on minimisation of the Akaike information criterion. The model was developed in R (v3.3.2, R Foundation, Vienna, Austria).

### Outcome

The primary outcome of this validation analysis was to identify the probability of an individual requiring a length of acute hospital stay following TJA equal to or greater than 5 days, or the probability of patients staying for the target 3 days or less. At present, current hospital processes rely on post operative factors to identify that a length of stay of greater than 5 days is imminent, at which time a transfer to an extended stay rehabilitation facility is initiated. The end of surgery date and time (hh:mm), and discharge date and time were extracted by data linkage to a surgical data electronic system within the hospital to calculate length of stay. For the purposes of model/tool assessment, the length of stay was recategorised based on the threshold of each original model/tool (**Table 1**).

### Participants/Records

The integrated electronic medical record (iEMR) was queried for all elective total hip and knee arthroplasties (no exclusions for revisions or bilateral surgeries) performed within the department between the dates listed. The query used the billing codes associated with total joint replacement for the hip and knee as dictated by the Australian Federal Medical Benefits Scheme (MBS items 49318 excluding trauma, 49319, 49324, 49327, 49330, 49333, 49345, 49346, 49518 and 49519). The results of the iEMR were made reidentifiable through removal of patient name, address, contact information. The query returned 1215 cases forming the database population **(Figure 1**). The available pool of records was limited to those that were raised following the hospital transition to iEMR in April 2019, leaving a pool of 331 patient records. Random permutation ^20^ between 1 and 200 was used to select the final list of 200 patients with a random number generator function, set to zero seeds (Matlab 2018b, Mathworks Inc, USA).

**Figure 1.**
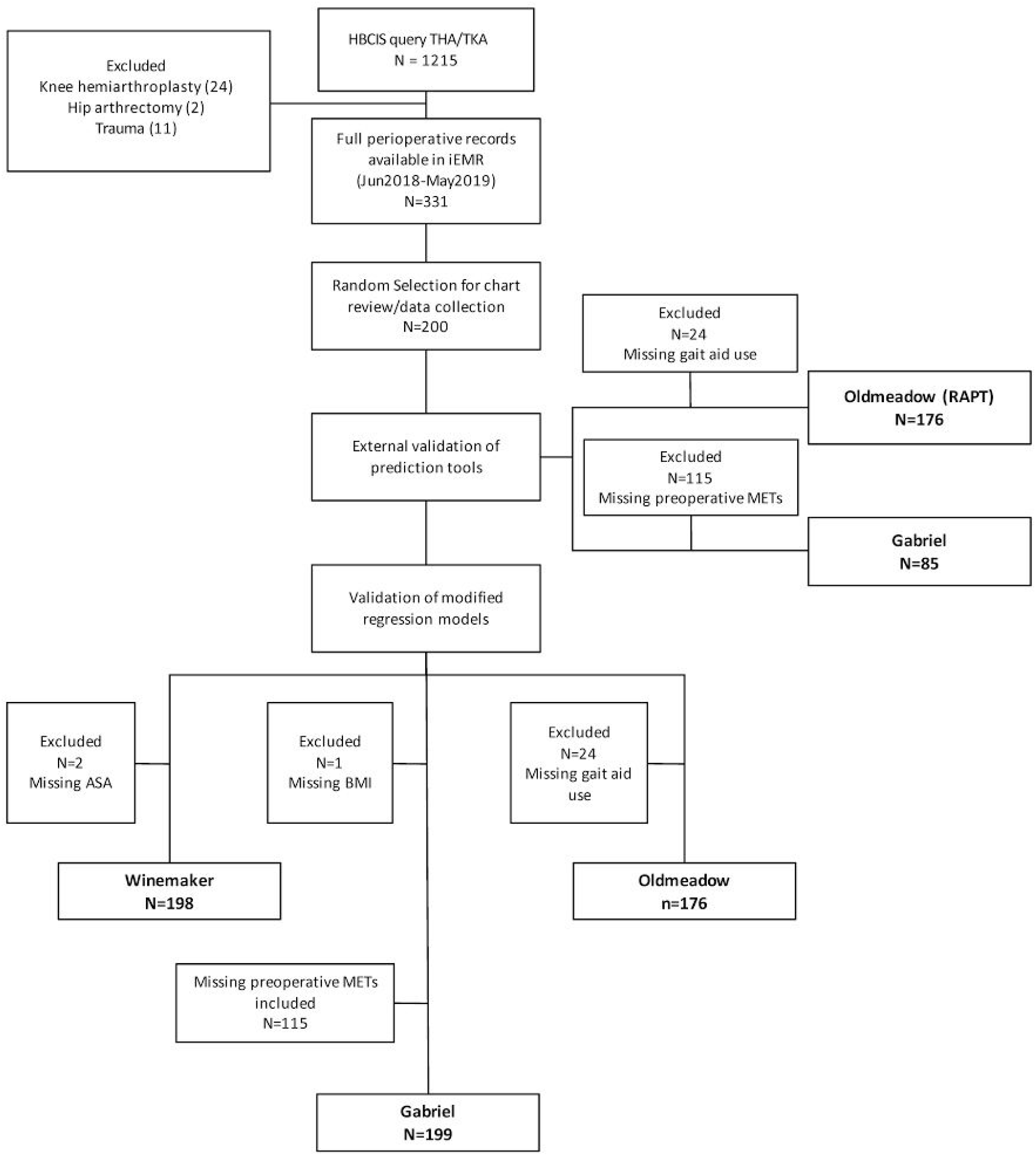
STROBE flow diagram from initial hospital record query to model/tool validation - 01-Jun-2018 to 04-Apr-2019

### Data sources, collection and linkage

Prior to data collection, a study core dataset was developed, combining all outcome and predictor variables from the selected models/tools as well as key indicators for patient data linkage (***Supplementary material A***). The research group comprised a collection team, led by orthopaedic physiotherapists and a quality team, based offsite to the hospital. The core dataset was agreed by both teams, and then converted to a master database for construction through data linkage, chart review, as well as variable calculations and recoding. Data linkage for eligible patients was performed between exports from different hospital systems provided by the hospital’s decision support unit using unique patient identifiers (unit record number; URN) for exact matches between datasets. Date and time of admission as well as discharge were extracted from the data linkage process, and used to calculate length of stay. Two reviewers performed a manual chart review of the patient’s electronic medical record for the remaining data items.

### Data quality

Data quality was assessed during the data collection period by export of the mastersheet weekly and imported into Matlab (v2018b, Mathworks Inc, USA) for further processing. A script was used to i) check for data completeness relative to the records marked *completed* by the collection team and ii) check that responses matched the core dataset framework, iii) check for outliers in the continuous variables and iv) assess consistency between variables for accurate patient inclusion/exclusions. Discrepancies were highlighted to the collection team through a dedicated column *(discrepancies)* in the mastersheet and confirmed with additional record review prior to model/tool adaptation and validation.

### Bias

Selection and information bias associated with data availability relative to the length of time from changeover to iEMR was mitigated by using random selection from the pool of all eligible records, rather than reverse chronological order.

### Modifications

At the conclusion of the data collection phase, with all data discrepancies resolved within the mastersheet, it was transferred to Matlab for further processing. The dataset was parsed into subsets reflecting each model and tool variable list. For external validation with the current dataset, the scoring tools and models included for assessment were modified to match the data available (**Supplementary material B**) and assumptions about the nature of the target dataset had to be made. The main reasons for adaptation were:

- Variable not routinely collected in the medical record
- Data not captured within the medical records in the form as specified in the model
- Variable included in the model was not able to be captured *preoperatively*
- Missing data at the individual record level

### Analysis and Model/tool validation

The final dataset (N = 200) was assessed for descriptive statistics on each of the variables included in at least one of the models/tools, and for missing data (***Supplementary material C***). Continuous variables were assumed to be non-normal and reported with median and interquartile ranges. Length of stay was visualised as a continuous variable and parsed into a three-level categorical variable (≤3 days; 4 days; ≥5 days). For the scoring tools (1 and 2) the predicted risk classification relative to the intended target category from the score was compared to the actual LoS data. Classification measures and confusion matrices were used to assess the performance of each tool to classify extended LoS. Matlab scripts (v2018b, Mathworks Inc, USA) were developed to run the adapted models using a generalized logistic regression model with intercept and linear structure to test main effects for the nominated predictors. No interactions were included and stepwise feature selection was not utilised. We assessed the predictive performance of the adapted models by examining measures of discrimination and calibration as oulined in the TRIPOD guidelines ^21^. The concordance index, identical to the area under the receiver operating characteristic curve (ROC) ^22^] was used to assess discrimination performance of the models. The goodness of fit between the predicted and observed risks was assessed using a calibration curve and Spiegelhalter’s Z-test ^23^ converted from an R script ^24^ to Matlab (Code Attachment), with an ideally calibrated model defined as one perfectly corresponding to the 45 degree reference line. Using the Z-test, model calibration is deemed inadequate if p_z_<0.05, and a perfectly calibrated model is defined as a Z score of zero, with larger deviations from zero corresponding to poorer calibration.

## Results

### Patient characteristics

A random selection of 200 patients was extracted for analysis, comprising a majority female and obese sample (**Table 2**). The majority of total joint arthroplasties (89%) were performed for osteoarthritis, with 83 hip procedures and 117 knee procedures. The majority of patients (70%) presented with at least one cardiovascular comorbidity (hypertension or atherosclerosis), with endocrine (47.5%) and gastrointestinal diagnoses (43%) also commonly noted.

The length of stay (LoS) ranged from 1.8 to 20 days at a median of 3.8 days (IQR 3 - 5), with 60% of patients discharged within 3 days of surgery, 16% of patients discharged at 4 days, and 24% at 5 days or greater. The majority (93%) of patients were discharged home with community support defined as in the care of relatives, spouses or carers; 5% required admission to inpatient rehabilitation, and 2% were discharged home to recover independently.

### Data coverage

‘Complete’ data were available in 176 patients for the adaptation of the RAPT tool ^2^, and 85 patients for the tool reported in Gabriel et al.^3^. Likewise, the three regression models were successfully adapted to include 198 (Winemaker) ^1^; 199 (Gabriel) ^3^ and 176 (Oldmeadow) ^2^ patients, respectively. Further information regarding missing data is available in ***Supplementary material C; tables C1 and C2***.

**Table 2.**
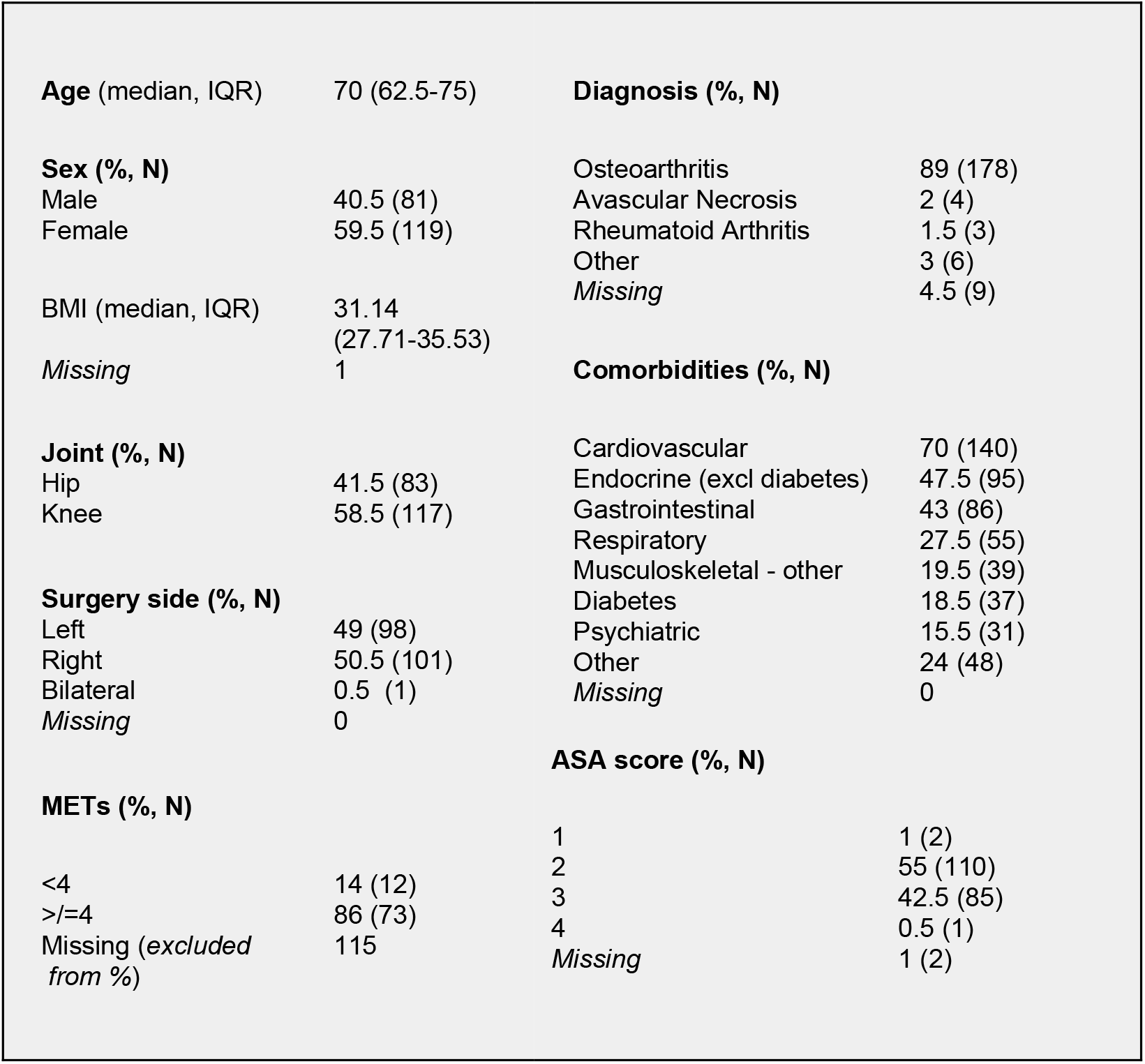
Demographics, Primary diagnosis, ASA scores, comorbidities and METS

|  |  |  |  |
| --- | --- | --- | --- |
| **Age (median, IQR)** | 70 (62.5-75) | **Diagnosis (% , N)** |  |
| **Sex (% , N)** |  | Osteoarthritis | 89 (178) |
| Male | 40.5 (81) | Avascular Necrosis | 2 (4) |
| Female | 59.5 (119) | Rheumatoid Arthritis | 1.5 (3) |
|  |  | Other | 3 (6) |
| BMI (median, IQR) | 31.14<br>(27.71-35.53) | Missing | 4.5 (9) |
| Missing | 1 | **Comorbidities (% , N)** |  |
| **Joint (% , N)** |  | Cardiovascular | 70 (140) |
| Hip | 41.5 (83) | Endocrine (excl diabetes) | 47.5 (95) |
| Knee | 58.5 (117) | Gastrointestinal | 43 (86) |
|  |  | Respiratory | 27.5 (55) |
| **Surgery side (% , N)** |  | Musculoskeletal - other | 19.5 (39) |
| Left | 49 (98) | Diabetes | 18.5 (37) |
| Right | 50.5 (101) | Psychiatric | 15.5 (31) |
| Bilateral | 0.5 (1) | Other | 24 (48) |
| Missing | 0 | Missing | 0 |
|  |  | **ASA score (% , N)** |  |
| **METs (% , N)** |  | 1 | 1 (2) |
| <4 | 14 (12) | 2 | 55 (110) |
| >=4 | 86 (73) | 3 | 42.5 (85) |
| Missing (excluded from %) | 115 | 4 | 0.5 (1) |
|  |  | Missing | 1 (2) |

### Predictive scoring tools

When applied to 176 eligible patients, the adaptation of the RAPT tool ^2^ accurately classified 36 of 42 patients who stayed 5 days or longer, however it also misclassified 91 of 134 patients who stayed for fewer than 5 days (**Figure 3**). This corresponds to a sensitivity of 85.71% (95%CI 71.46-94.57), and specificity of 32.09% (24.29-40.70; **Table 3**). Of the limited sample (N=85) available for validation of the second scoring tool ^3^, 41 of 60 patients who stayed longer than 3 days, and 11 of 25 patients who stayed 3 or fewer days were correctly classified, corresponding to a sensitivity of 44% (24.4-65.07) and specificity of 68.33% (55.04-79.74), respectively. When used to classify patients against the target threshold of 5 or more days, the second tool correctly classified 11 of the 15 patients with an extended stay, however it misclassified 44 of 70 patients who stayed fewer than 5 days.

**Table 3.** External Validation of Prediction Tools.

|  | Oldmeadow et al. (2003) ≥5 d |  | Gabriel et al. (2018) ≥5 d |  | Gabriel et al. (2018) > 3 d |  |
| --- | --- | --- | --- | --- | --- | --- |
|  | Value | 95%CI | Value | 95%CI | Value | 95%CI |
| *Sensitivity (% 5+d or 3+d correctly identified)* | 85.71% | 71.46% to 94.57% | 73.33% | 44.90% to 92.21% | 44.00% | 24.40% to 65.07% |
| *Specificity (% <5d or <3d correctly identified)* | 32.09% | 24.29% to 40.70% | 37.14% | 25.89% to 49.52% | 68.33% | 55.04% to 79.74% |
| *Positive likelihood ratio* | 1.26 | 1.07 to 1.50 | 1.17 | 0.82 to 1.66 | 1.39 | 0.78 to 2.48 |
| *Negative likelihood ratio* | 0.45 | 0.20 to 0.97 | 0.72 | 0.29 to 1.75 | 0.82 | 0.56 to 1.21 |
| *Estimated prevalence* | 23.86% | 17.77% to 30.86% | 17.65% | 10.23% to 27.43% | 29.41% | 20.02% to 40.29% |
| *PPV* | 28.35% | 25.03% to 31.92% | 26.92% | 20.54% to 34.43% | 67.58% | 53.91% to 78.79% |
| *NPV* | 87.76% | 76.65% to 93.99% | 81.52% | 64.36% to 91.50% | 44.86% | 35.57% to 54.52% |
| *Accuracy* | 44.89% | 37.40% to 52.55% | 45.83% | 34.97% to 56.99% | 53.73% | 42.58% to 64.62% |

**Figure 3.**
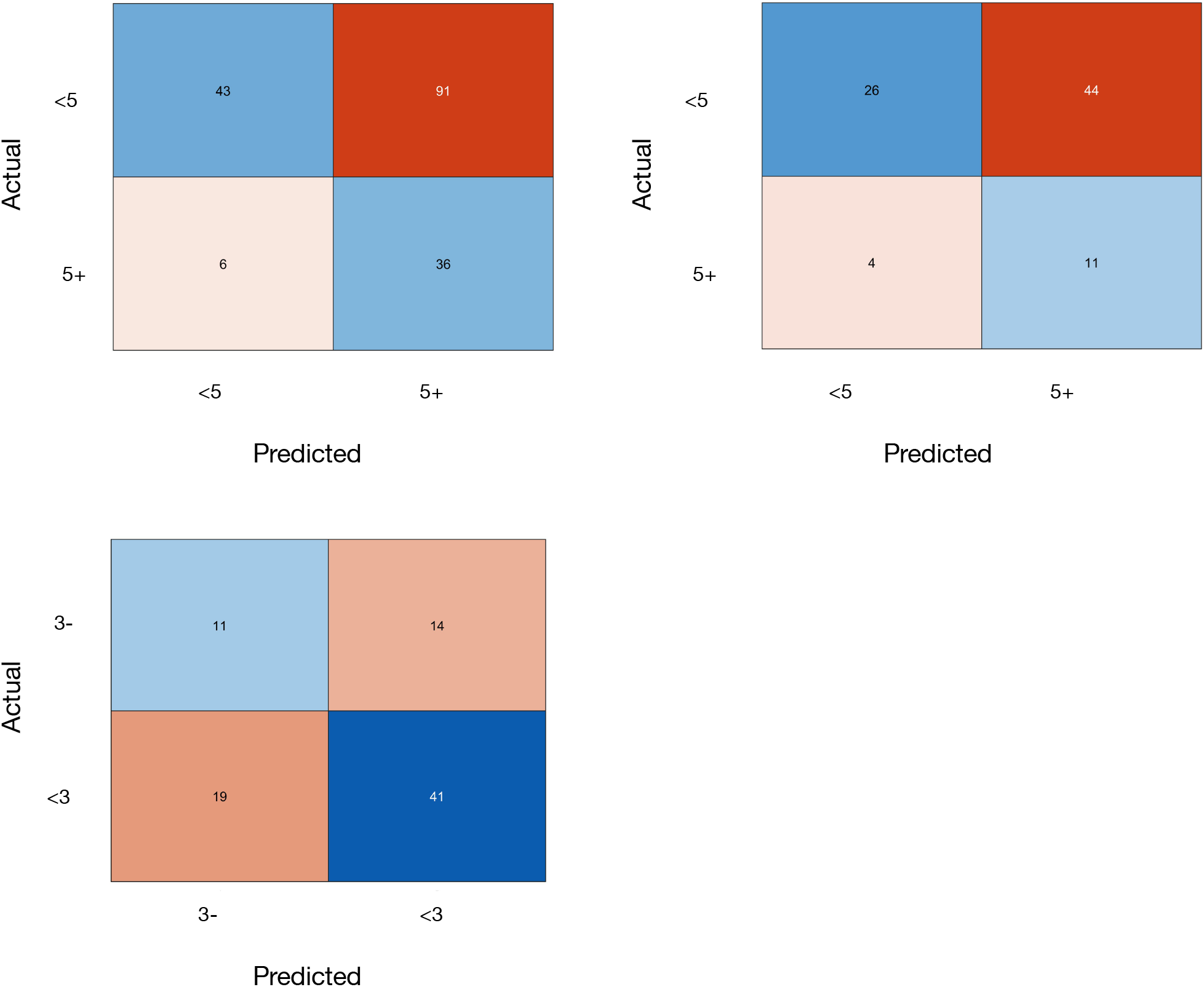
Confusion Matrices for Predictive Tool Performance, Predicted length of stay vs actual length of stay. Top Left, Tool 12; Top Right, Tool 23 score validation extended stay (≥5 days); Bottom Left, Tool 2 score validation target stay (3-days).

### Predictive regression

#### models Model results

Adapting the model described by Winemaker et al to the study dataset, four key independent predictive factors were identified; patients 75 years or over (OR 8.85; p<0.001) and those undergoing THA as opposed to TKA (OR 2.69; p=0.03) were more likely to fall into the extended stay (≥5 days) category, with males (OR 0.33; p<0.001) and patients with renal comorbidities (OR 0.15; p=0.05) less likely to experience an extended stay. Using the adaptation of the model described by Oldmeadow et al, three key protective factors were identified; male patients (OR 0.3; p=0.008), those aged 50-65 (OR 0.18; p=0.002), and those aged 66-75 (OR 0.2; p<0.001) were less likely to stay 5 days or longer. For the adaptation of the model described in Gabriel et al, only male gender (OR 2.59; p=0.005) was predictive of a hospital stay of 3 days or less.

#### Model performance

Discrimination performance of the three models was similar, with the concordance index ranging between 0.7 and 0.79, similar to the values of 0.73 reported for the original model by Winemaker et al., and 0.76 (95%CI 0.7-0.811) for the model by Gabriel et al. Calibration testing did not identify any of the three models as poorly calibrated to the data (Spiegelhalter p>0.05). Visual inspection of calibration indicated a degree of inaccuracy most evident in patients stratified into the intermediate risk regions of the two models designed to identify potential cases staying 5 days or longer.

**Figure 4.**
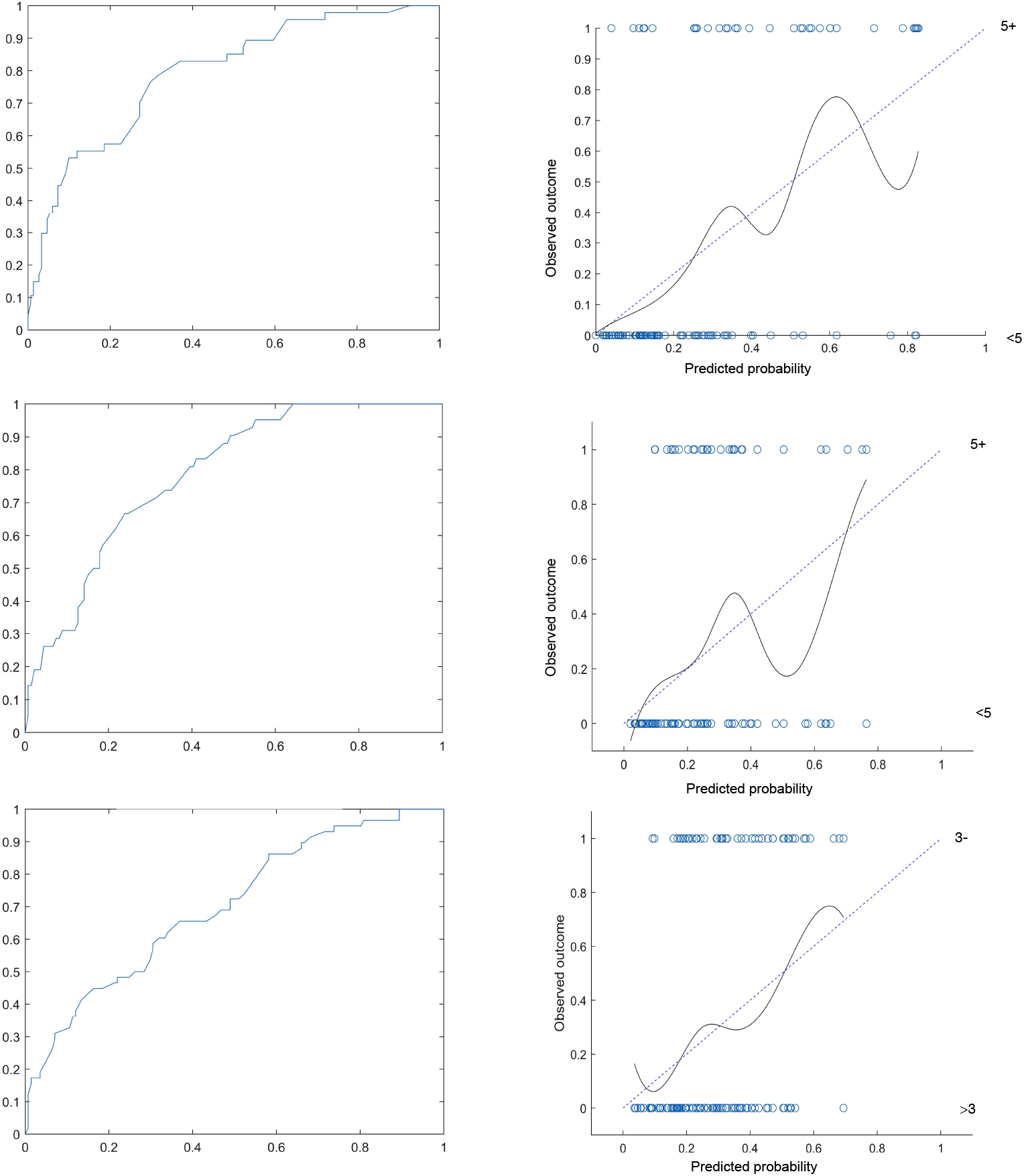
Receiver operating curve and calibration curves of adapted regression models. *(Top)* Winemaker et al., 2015 - (N=198) *(Middle)* Oldmeadow et al., 2003 (N=176). *(Bottom)* Gabriel et al., 2018 [less than or = 3 day cutoff] (N=199).

**Table 3.**
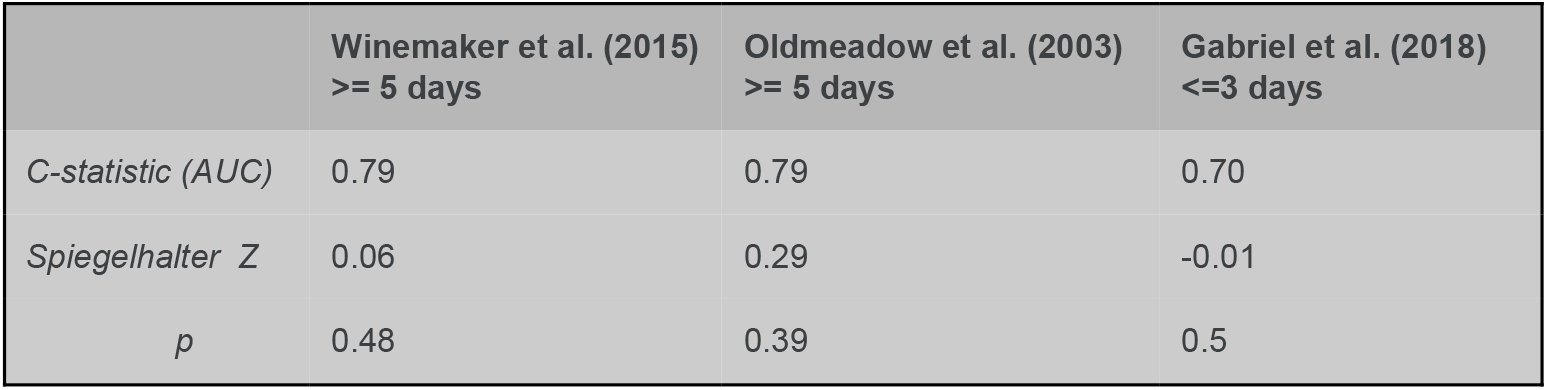
Performance metrics of regression models

|  | Winemaker et al. (2015)<br>≥ 5 days | Oldmeadow et al. (2003)<br>≥ 5 days | Gabriel et al. (2018)<br>≤ 3 days |
| --- | --- | --- | --- |
| *C-statistic (AUC)* | 0.79 | 0.79 | 0.70 |
| *Spiegelhalter Z* | 0.06 | 0.29 | -0.01 |
| *p* | 0.48 | 0.39 | 0.5 |

## Discussion

### Recap

Predictive model development for hospital decision-making is dependent on sufficient data linkage, and a sufficient volume of patients to reach adequate sample sizes for development and internal validation ^22^. Thus smaller sites run the risk of developing models that quickly become obsolete due to changes in procedure or patient characteristics over time. Thus the present study represents a pragmatic strategy for assessing the performance of three previously existing predictive models after adaptation to an independent hospital site for the purpose of identifying patients at risk of an extended hospital stay (≥5 days) following lower limb arthroplasty. The eventual clinical utility of this model is to select patients most likely to benefit from changes in hospital procedure, with the end goal of enabling earlier allocation of resources to patients likely to require an extended stay in hospital following their joint replacement surgery.

### Tool evaluation

A direct application of the prediction tools (with modifications based on available data) reported in Oldmeadow (RAPT) and Gabriel were not successful, with large misclassification errors preventing reliable identification of patients at risk of extended stay, or those likely to leave within a more ideal 3 day time frame.

External validation of prediction tools developed at other sites suffers from a number of major challenges, a number of which were observed here. Firstly, any particularly unique hospital characteristics, such as patient demographics and procedures, likely impact the typical length of stay following hip or knee arthroplasty. This means that factors identified elsewhere may not impart the same predictive value at a new site. One particularly salient example is the discrepancy in discharge destination between the Oldmeadow patient cohort and the present cohort; while 95% of patients were discharged home with access to community support services, 44.5% of the patients in the Oldmeadow study were discharged to extended inpatient rehabilitation. This most likely reflects a move toward short-stay protocols occurring over the last ten years or so, as well as evolutions in surgical procedures and postoperative management (e.g. physiotherapy) leading to improved patient recovery in the early days following surgery.

Secondly, a number of items listed in the tools were not routinely captured in the patients’ electronic medical records. In particular, patient-reported factors including availability of community support, walking distance and use of gait aids, and opioid dosage were either absent or missing from many electronic records. Given a lack of community support (ie. family or carer help) is a well-known barrier to discharge for patients undergoing knee and hip replacement ^5,6,25^, any attempts at prospectively identifying patients at risk of an extended hospital stay should ensure psychosocial variables such as this are taken into account.

### Regression model performance

All three regression models appeared to be adequately calibrated to the available retrospective hospital data, and a concordance index between 0.7-0.8 indicated they all possessed acceptable discriminatory performance, similar to the original published models. The coverage of data and performance of the models based on those by Winemaker et al^1^ and Gabriel et al^3^ in particular appear to indicate clinical utility for selection of patients to target for improvements in length of hospital stay.

However, not all risk factors identified in the previous models could be replicated in the present dataset. This may largely be due to sample size constraints imposed by the source of data, which contained records restricted to a one-year period from a medium-sized metropolitan hospital. In particular, the adaptation of the tool described by Gabriel et al. was characterised by a substantial amount of missing data, which may explain the failure to identify any independent risk (or protective) factors besides gender. Nonetheless, when the underlying model was adapted to the available dataset by including "not reported” as a response category for METS, it demonstrated adequate performance metrics in the present cohort, suggesting the remaining items still have clinical utility. While point estimates of calibration suggest all three models were not poorly calibrated to the data, a degree of inaccuracy is evident in each model, particularly amongst patients with ‘intermediate’ risk profiles. This is consistent with similar studies, suggesting key risk factors have not been accounted for ^2,26^. It is likely that in patients with a small number of comorbidities, patient-reported psychosocial or functional risk factors are crucial for differentiating those who are likely to leave hospital within the target time frame from those who stay longer ^2,27^. Depending on the resourcing requirements, it would be advisable to either 1) incorporate routine capture of this data into the electronic medical record to enable more predictive accuracy in this subset of patients, and/or 2) ensure preoperative planning addresses psychosocial factors including patient expectations of discharge, psychological health, and engagement with early postoperative rehabilitation.

### Limitations

Overall, a few factors limited the ability of this study to confidently validate the chosen models in a new hospital setting. First, the availability of data was limited in date to a single year due to changeover in electronic hospital systems. Additionally, the scope of the available data did not fully match the reference models. This limited the sample size available for assessment. While hospital-level data is unlikely to be detailed enough (and may not be necessary) to capture all relevant factors affecting a patient’s length of stay, a few notable omissions are worth mentioning. Firstly, revision surgeries (especially those performed for periprosthetic joint infection) are associated with greater risk of complications leading to an extended hospital stay ^15,28,29^. Although revision surgeries made up only 3.5% of cases in the present case mix (and 4.2% of extended stay cases), omitting the type of surgery from a predictive model may lead to a degree of inaccuracy. Secondly, classification of certain comorbidities may impact their accuracy in predicting extended stay. For example, cardiovascular comorbidities incorporate conditions which are relatively simple to manage (eg. hypertension), as well as those likely to require additional procedures to manage risk during surgery, such as atrial fibrillation. In the context of limited population data, predictive modelling may not require individual patient-level precision in order to be clinically useful ^30^, and such tools do not replace the patient-level decision making which takes place prior to surgery. Therefore, selection of the ideal predictive model for clinical adaptation requires a detailed consideration of how it will integrate into the clinical decision-making process.

## Conclusion

This study reports an assessment of three regression models intended to identify patients at risk of an extended hospital stay after total hip or knee arthroplasty. While an adaptation of the two prediction tools reported previously performed poorly on the present dataset, the three regression underlying models performed adequately in terms of discrimination and calibration. Although model performance was similar, it appears the models based on Winemaker et al. ^1^ and Gabriel et al. ^3^ had attributes that indicated potential for successful application within the clinical setting, with adequate coverage of relevant data in the iEMR and good performance of the Winemaker model, and good calibration and acceptable discrimination performance of the Gabriel model in spite of missing METs values. (ie. existing coverage of data in iEMR, or good calibration in spite of missing data). Further evaluation of the resourcing implications of the selected models and their target groups is required in order to optimise data capture and targeting of resources to patients at risk of an extended hospital stay, which is expected to enhance patient outcomes overall and reduce the burden on hospital resources.

## Data Availability

Data may be available from the corresponding author upon reasonable request.

## Acknowledgements

Thanks to the nursing and allied health staff at the Queen Elizabeth II Jubilee Hospital, Brisbane, Qld Australia and the data analysis staff at EBMA Analytics, Sydney, NSW Australia for support and assistance with this study. The authors acknowledge the assistance of Kerry Baldwin (Physiotherapy) and Maha Jegatheesan (Orthopaedics) for their contributions to chart review and Thomas Cooper (EBMA) for his contributions to the analysis code.

## Supplementary material

**Supplementary material A: Variable definitions**

**Supplementary material B: Adaptation of models and tools for validation against the current dataset**

**Supplementary material C: Missing data by variable**

**Supplementary material D: Code attachment**

